# Deep learning based quantification of the accelerated brain aging rate in glioma patients after radiotherapy

**DOI:** 10.1101/2021.12.22.21267899

**Authors:** Selena I. Huisman, Arthur T.J. van der Boog, Fia Cialdella, Joost J.C. Verhoeff, Szabolcs David

**Affiliations:** Department of Radiation Oncology, UMC Utrecht, 3584 CX Utrecht, The Netherlands; Department of Medical Oncology, UMC Utrecht, 3584 CX Utrecht, The Netherlands

**Keywords:** Brain age prediction, Deep learning, Glioma, Radiation Therapy

## Abstract

**Background and purpose:** Changes of healthy appearing brain tissue after radiotherapy have been previously observed, however, they remain difficult to quantify. Due to these changes, patients undergoing radiotherapy may have a higher risk of cognitive decline, leading to a reduced quality of life. The experienced tissue atrophy is similar to the effects of normal aging in healthy individuals. We propose a new way to quantify tissue changes after cranial RT as accelerated brain aging using the BrainAGE framework.

**Materials and methods:** BrainAGE was applied to longitudinal MRI scans of 32 glioma patients, who have undergone radiotherapy. Utilizing a pre-trained deep learning model, brain age is estimated for all patients’ pre-radiotherapy planning and follow-up MRI scans to get a quantification of the changes occurring in the brain over time. Saliency maps were extracted from the model to spatially identify which areas of the brain the deep learning model weighs highest for predicting age. The predicted ages from the deep learning model were used in a linear mixed effects model to quantity aging and aging rates for patients after radiotherapy.

**Results:** The linear mixed effects model resulted in an accelerated aging rate of 2.78 years per year, a significant increase over a normal aging rate of 1 (p *<* 0.05, confidence interval (CI) = 2.54-3.02). Furthermore, the saliency maps showed numerous anatomically well-defined areas, e.g.: Heschl’s gyrus among others, determined by the model as important for brain age prediction.

**Conclusion:** We found that patients undergoing radiotherapy are affected by significant radiation-induced accelerated aging, with several anatomically well-defined areas contributing to this aging. The estimated brain age could provide a method for quantifying quality of life post-radiotherapy.

**Highlights:** Up to 3 times accelerated aging after radiotherapy. // Anatomically well-defined areas for brain age prediction. // Quantifying quality of life after radiotherapy.

## 1 Introduction

One of the primary treatments for tumors in the brain is radiation therapy (RT), usually with a combination of surgery, chemotherapy [1, 2] and, occasionally, immunotherapy [3]. Due to the complex nature of changes in the brain, quantifying the effects of RT on seemingly healthy brain tissue remains a challenge. Tissue atrophy occurs both post-RT and in normal aging, with atrophy caused by normal aging occurring at a low rate of ∼0.5% per year in the healthy elderly [4]. As tissue atrophy caused by RT resembles accelerated natural aging, the ability to calculate a patient’s ”brain age” to quantify atrophy could provide useful insights due to ease of interpretation compared to other methods such as cortical thickness and volumetric measures, which rely on pre-engineered features [5–7]. One method to quantify brain age is the BrainAGE framework [8], which has already been widely used in describing disease-related changes of the brain, such as Alzheimer’s disease [8, 9] and psychiatric disorders [10, 11]. BrainAGE, or brain age gap estimation, is a technique to determine the discrepancy between a person’s chronological- and biological brain age [12], using magnetic resonance imaging (MRI) scans. In a healthy brain, showing normal aging patterns, the chronological and biological ages are expected to be identical, however, in case of an abnormal condition or disease the biological age may differ from the chronological age. In this study, BrainAGE will be applied in glioma patients who have undergone RT.

RT plays an important role in the treatment of cranial tumors, however, the effect of radiation is not selective to cancer affected tissue and it comes with the unintentional side effect of radiation-induced brain injury to the rest of the brain tissue, which can lead to progressive cognitive decline. Cognitive symptoms occur in approximately 50-90% of patients undergoing RT [13], and can lead to a reduced quality of life (QoL) [14]. Quantifying changes in the brain using an age-based metric is of interest, as it can be related to some of the changes in QoL using existing knowledge on brain aging [15]. By predicting a brain age of a patient before RT and comparing that age to the ages predicted for the follow up scans, the effects of RT on brain aging can be determined in a longitudinal manner. Since the effects of RT on cancer-affected tissue are extremely destructive, a similar effect can be expected on the healthy tissue. Since the effects of RT on the healthy tissue are similar to aging in a healthy brain, understanding age-related changes in a healthy brain is of importance. Two examples of healthy brain aging is shown in figure 1. In figure 1/A, an example of aging over a longer time span in cross-sectional data is shown from the Information eXtraction from Images (IXI) data set [16]. Noticeable differences are in the size of the ventricles and space between the gyri, indicating a loss of tissue volume. An example of relatively short-term aging is shown in figure 1/B, taken from the MyConnectome data set [17]. The time between the scans is approximately 11 months, which is similar to the mean follow-up time of 11.67 months between first and last scan within our RT data set. The short-term (non-)aging example shows that the healthy brain is not affected by tissue atrophy in a similar time span as the expected overall survival of glioma patients. The differences between the scans, most notably the slightly darker colour of the brain stem in the second scan, are related to the different noise patterns and contrast.

**Fig. 1.**
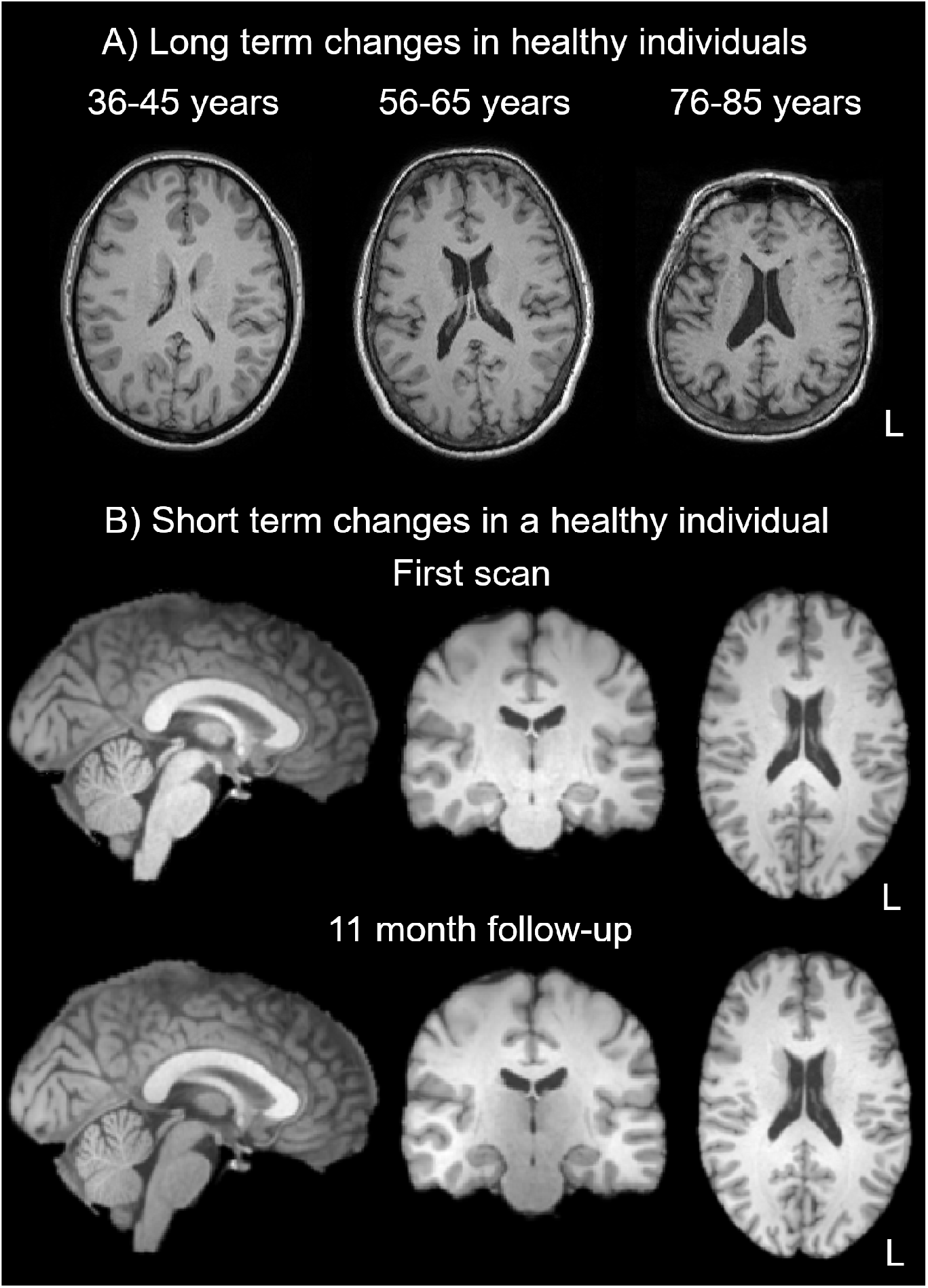
Two examples of aging, with figure 1/A showing long term aging in cross-sectional cohort from the IXI data set [16], showing 3 MRI scans for the ranges of 36-45 years, 56-65 years and 76-85 years, from left to right respectively. Figure 1/B shows two longitudinal MRI scans from the MyConnectome project [17], taken approximately 11 months apart. Both MyConnectome scans were preprocessed using optiBET for brain extraction [18] and FLIRT for linear registration to MNI152 template. [19, 20]

## 2 Materials and Methods

### 2.1 Patient Selection and Data Collection

For this study, MRI scans of 32 histologically proven glioma patients, who received RT in the UMC Utrecht in the period from 2016 to 2017, were analyzed retrospectively. The first scan of every patient was acquired during the postoperative RT planning. The scans are T1-weighted and were acquiring on a 3T Philips Ingenia scanner with the 3D turbo-spin echo sequence without gadolinium enhancements. The voxel resolution is 1 × 0.96 × 0.96 mm^3^, with a matrix size of 207 × 289 and 213 continuous axial slices without gap. The parameters used were TR = 8.1 ms, TE = 3.7 ms and the flip angle was 8 degrees. The minimum number of scans per patient was 2, the pre-RT scan and the first scan after RT. The maximum amount of scans acquired was 9, with a mean of 4.03, and an SD of 1.96. The mean time between first and last scan is 11.67 months, with a SD of 4.29 months. See figure 2 for a visualization of the time between scans, and table 1 for an overview of patient characteristics. The institutional review board waived informed consent for this retrospective study (study ID 18/274). The IXI data set, utilized for validating the saliency maps was adjusted for this study to contain 310 healthy individuals between the age range of 42 and 82. This selection of individuals contains 134 males and 176 females, with a mean age of 59.19 and an SD of 9.27.

**Fig. 2.**
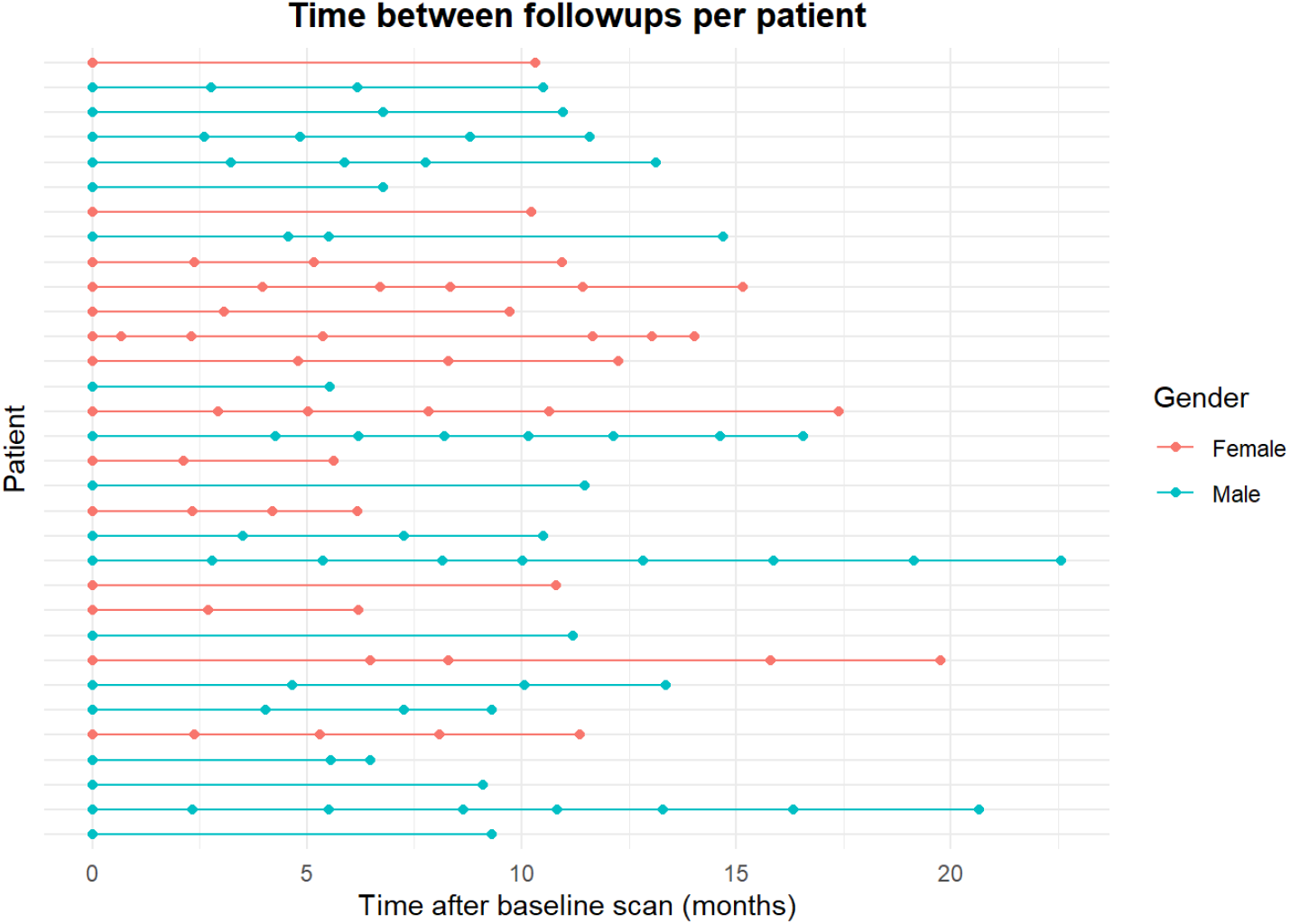
Visualization of the time between scans (x axis) per patient (y axis). Each dot represents a scan for a patient, with each patient starting at the pre-RT scan.

**Table 1.** Table of baseline patient characteristics.

|  | **N (total n = 32)** |
| --- | --- |
| Age | $57.64 \pm 9.01$ |
| Sex |  |
| Male | 18 (56.25%) |
| Female | 14 (43.75%) |
| WHO Grade |  |
| II | 10 (31.25%) |
| III | 3 (9.375%) |
| IV | 19 (59.375%) |

### 2.2 Preprocessing

To be able to utilize the model from Peng et al. [21] the data required preprocessing. Specifically, the data had to be reoriented to stereotaxic 1mm Montreal Neurological Institute (MNI) space and non-brain tissue had to be removed. Both of these actions were performed using FSL version 6.0. [22] First, the non-brain tissue was removed using *optiBET* [18], an optimized version of *BET* [23], using the default settings. The rigid registration to MNI space was done using the *FSL FLIRT* tool [19, 20], with 6 degrees of freedom, using the image output from *optiBET* and the MNI152 template as reference. Finally, the transformed images were subject to the pre-trained network to obtain a predicted age for each brain. In figure 3, a visualization of the complete pre-processing pipeline can be found. The MRI scan in this figure is from a patient between 41 and 45 years old, which will function as an example throughout the manuscript.

**Fig. 3.**
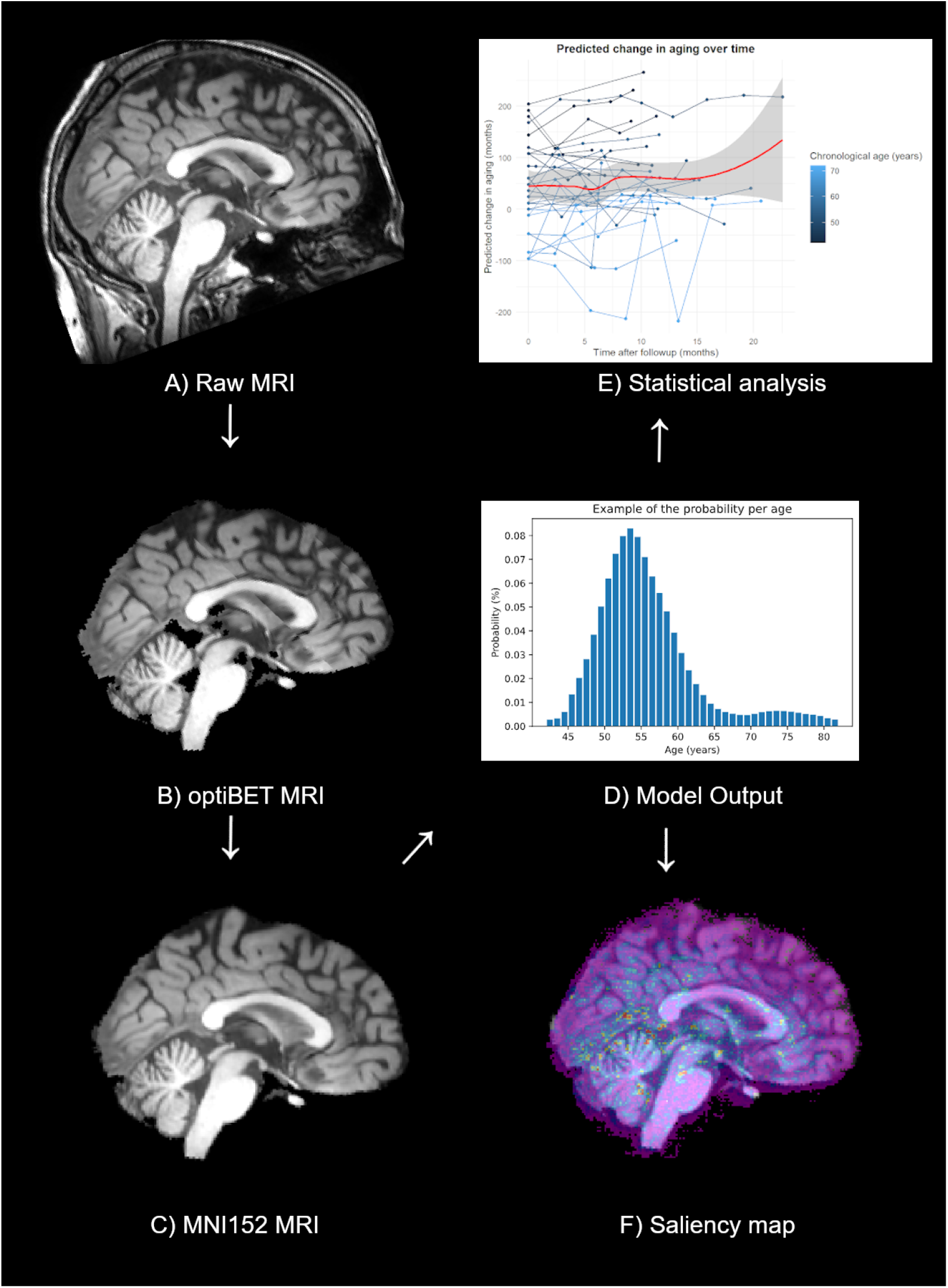
The processing pipelipe for applying the model, starting with A), the unprocessed MRI from the patient between 41 and 45 years old. The unprocessed MRI is processed using *optiBET* to obtain B), the *optiBET* MRI, which is then put into MNI152 space to get C), the MNI152 MRI. This MRI can then be used in the model, providing D), the model output, which can be used to perform E), the statistical analysis and obtain F), the saliency maps.

### 2.3 Deep learning model

The Simple Fully Convolution Network (SFCN) model by Peng et al. [21], which was trained on UK Biobank data [24] and selected for winning the PAC2019 contest [25], was used via *Python* 3.85 [26] to obtain a probability distribution for each of the MRI scans. This distribution ranged from ages 42 to 82, adding up to a total of 40 possible outcomes. The age with the highest probability was selected as the predicted age. For an example of the model output see figure 4, showing the histogram of the 41-45 year old patient pre-RT and the predicted age of 54.

**Fig. 4.**
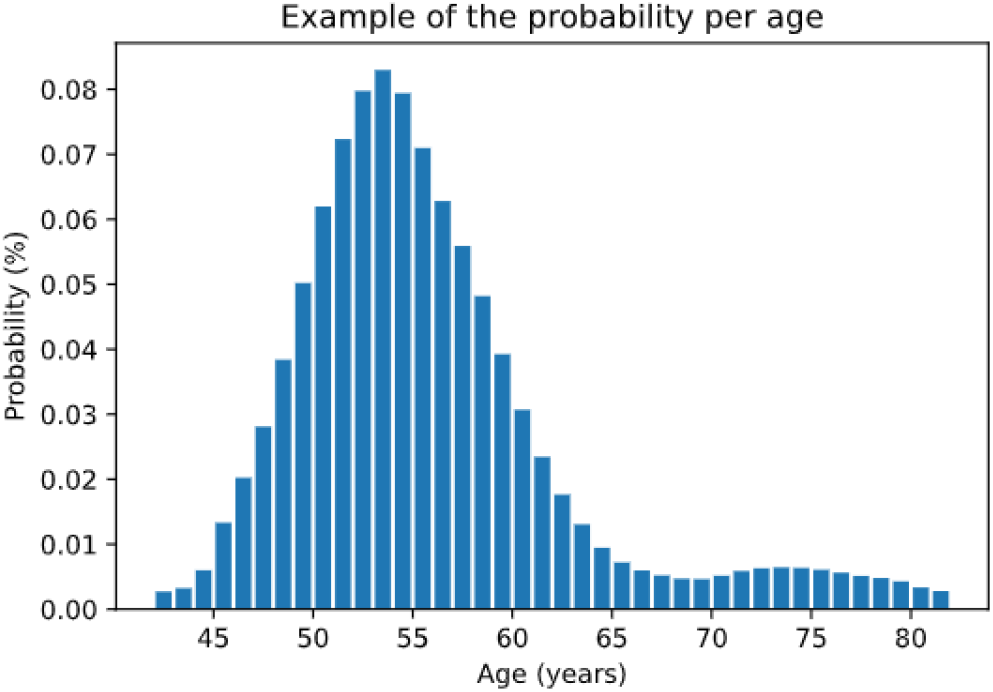
Probability distribution for the pre-RT scan from the 41-45 year old patient, with the x axis representing predicted age and the y axis representing the predicted probability of that age.

### 2.4 Saliency maps

Furthermore, saliency maps were extracted from the SFCN model to investigate which parts of the brain contribute to the age estimation the most. These saliency maps were created and averaged for both the RT data set and the IXI data set. By retrieving the voxel weights the model used to predict the ages for each patient and average them across the whole cohort, a visualization of the contribution of all brain areas was created. The FSL tool *autoaq* was used to aid the anatomical interpretation of such areas using built-in atlases [27–30]. An arbitrary threshold of 0.05 was selected to eliminate the voxels with a relatively low weight to aid visual interpretation and highlight hotspots. Finally, the saliency maps for both data sets were compared by subtracting the saliency maps from each other and removing all data points with a difference less than 0.015.

### 2.5 Statistical modeling

To obtain the aging rate for each patient, *RStudio* 1.2.5019 [31] with the package *‘lme4’* [32] was used to implement mixed models. For the linear mixed model, the formula

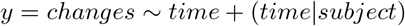

was used, where the ”changes” are the biological changes in aging in months, and the ”time” is the time passed in months. The model is adjusted for normal aging, so only accelerated or decelerated aging are predicted, with 0 being normal aging. The model predicts an age, using mixed effects linear regression for each patient, with the subject being an exclusively random effect, while the time passed is both a fixed and a random effect. By using ”time” as both a fixed and random effect, the average aging rate is used as predictor due to the fixed effect, and the random effect allows the aging rate to vary between patients. To correct for the baseline prediction error, the error was removed from all scans, taking the initial error per patient and subtracting it from every scan for that patient.

## 3 Results

### 3.1 Pre-RT scans

To test the accuracy of the SFCN model before RT, the pre-RT scans were first analyzed separately. The mean absolute error (MAE) for the pre-RT scans is 6.53 years, which is significantly higher than the 2.14 years found by Peng et al. [21]. In figure 5, the chronological age is compared to the predicted age. The figure shows that the ages of older patients are underestimated by the model, while the ages of younger patients are overestimated.

**Fig. 5.**
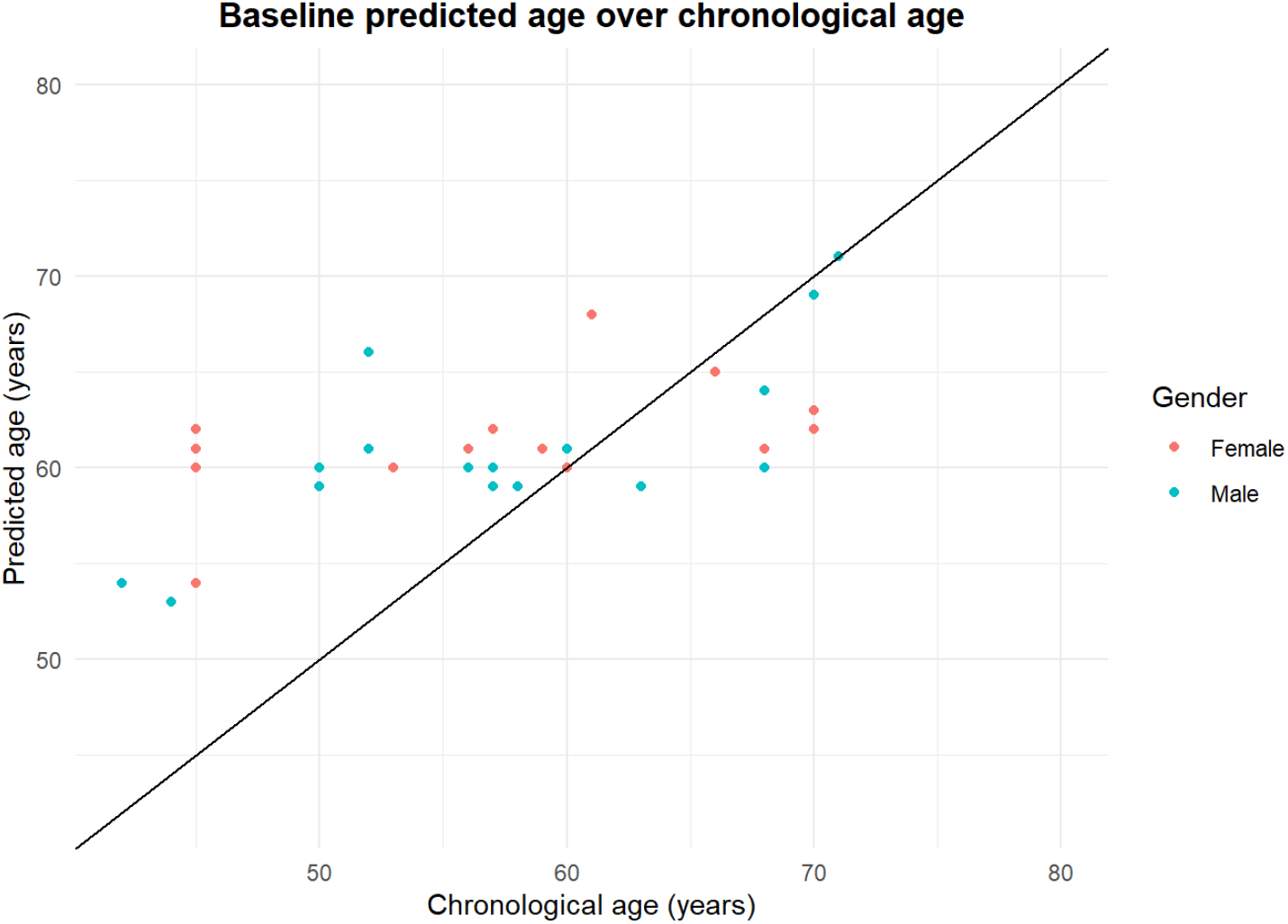
The age predicted for the pre-RT scans (y axis) compared to the chronological age (x axis) of the patient at the pre-RT scan. The black line shows y = x, where chronological age and biological age are the same.

### 3.2 Model output

Figure 6 shows three follow-up scans of the aforementioned 41-45 year old patient. The chronological age for this patient is between 41 and 45, while the SFCN model predicts 54 years for the pre-RT scan. Four months after RT at the first follow-up, the scan is predicted at 59 years, indicating a five year increase in biological age, or a BrainAGE score of +5 years in four months, which indicates a 15x aging rate. Similar effects are found based on the next two follow-up scans, showing that this particular patient’s brain aged a total of 8 years in the 9 months after RT. The clear upward aging trend presented for this patient is visible in the tissue changes.

**Fig. 6.**
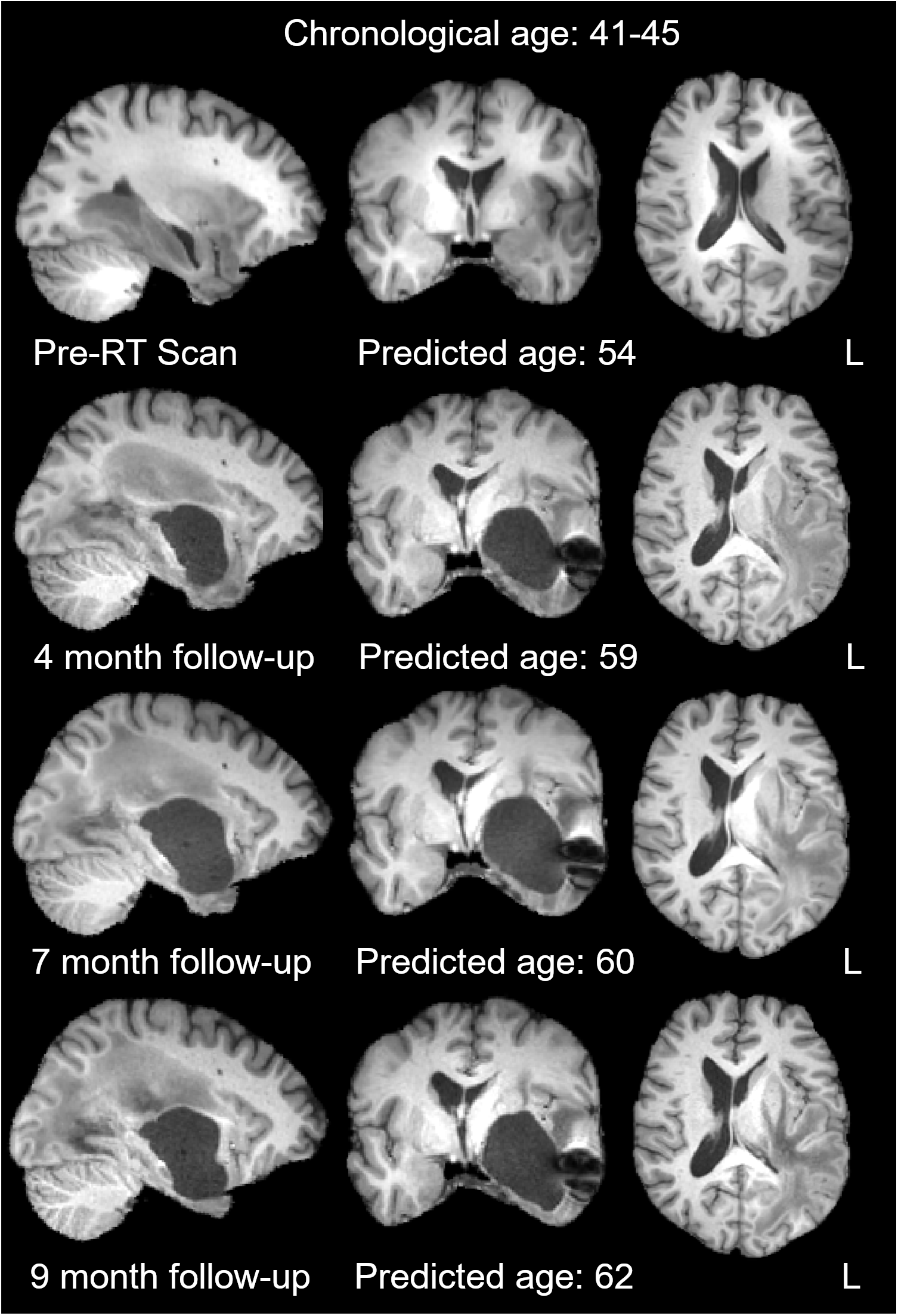
Four MRI scans from the same 41 to 45 year old patient, with the axial plane showing the ventricles, while the sagittal and coronal plane show the location of the tumor. The age is predicted by the SFCN model at pre-RT, and at each of the follow-ups.

### 3.3 Predicted aging

To analyze the changes in aging rate, the SFCN model predicted the ages for all images. Figure 7/A shows the predicted age for all scans, minus the chronological age (adjusted for normal aging). The red curve represents a smoothed average of all predictions using a locally estimated scatterplot smoothing (LOESS) function [33].

**Fig. 7.**
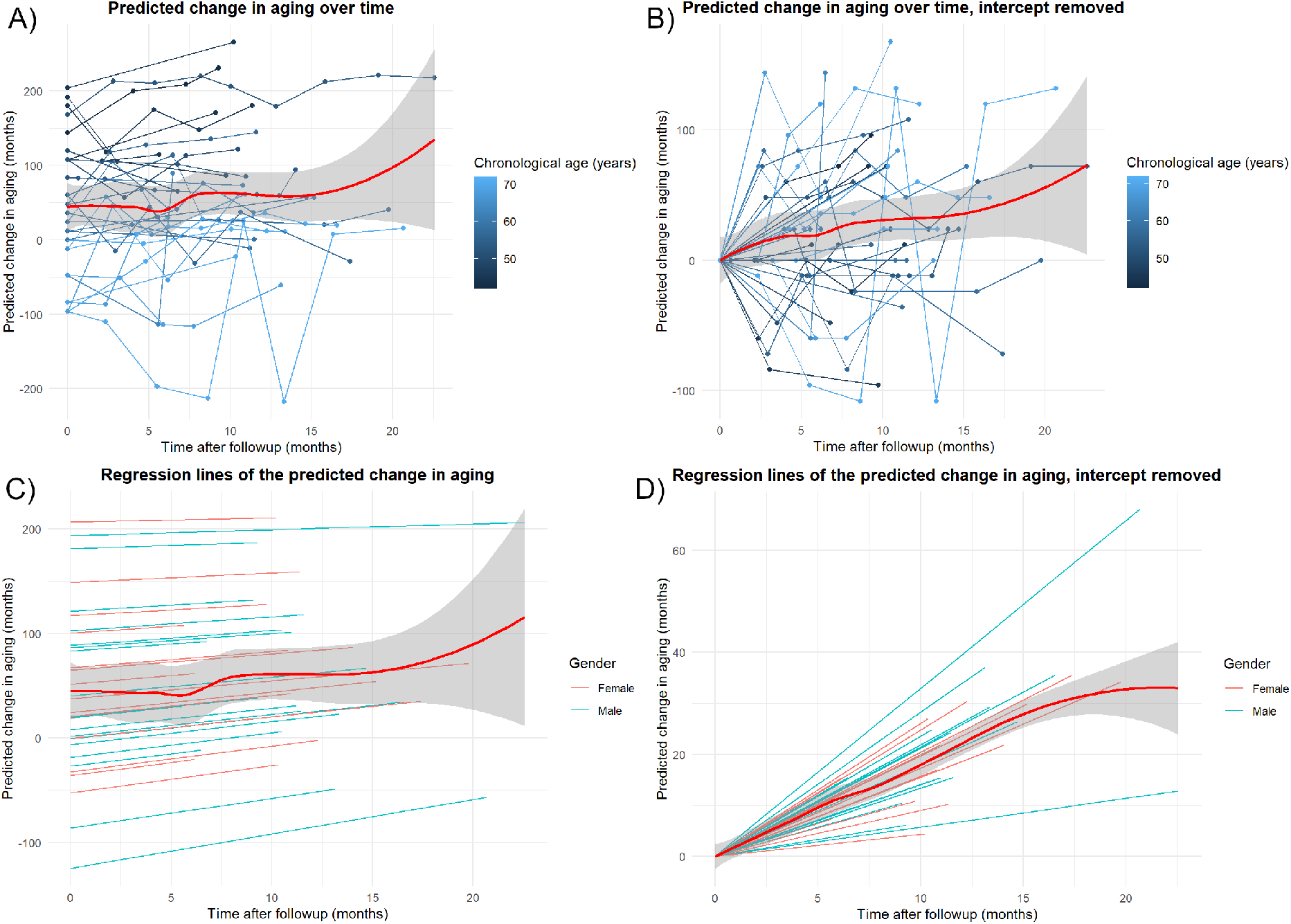
For each graph, the predicted change in aging (y axis) is shown over the time during follow-up in months (x axis). The red curves show the LOESS averaged regression curves, with the grey bands showing the 95% confidence interval for each respective curve. Each line represent a single patient, the points on the lines represent an MRI scan, and the horizontal line shows normal aging. Figure 7/A shows each of the predicted ages per scan per patient. Figure 7/B shows the same predicted ages, but adjusted for the baseline prediction error. For figure 7/A and 7/B the colouring is lighter blue for higher chronological ages, and darker blue for lower chronological ages. Figure 7/C shows the regression slopes prediction by the linear mixed effects model. Figure 7/D contains the same regression slopes with the intercept removed.

As this curve trends upward, an increased aging rate is implied as time passes. However, this analysis does not take into account the prediction error, which was shown based-on the pre-RT scans, resulting in a biased average. The averaged curve continues to trend upward towards the late follow-ups at the two year mark. The grey-coloured bands show that the confidence interval is much wider for this area, as the available data is more sparse in this time period. The confidence bands remain narrow in the first 12 months. Figure 7/B represents the predicted aging over time, corrected for the baseline prediction error. The smoothed average LOESS curve shows a positive slope and narrow confidence bands after the bias is removed. This indicates that the changes in aging, on average, are accelerated.

In figure 7/C, the individual aging rates per patient, as predicted by the mixed effects model based on the data in figure 7/A, are shown. The average predicted aging rate for all patients was 2.78, which is statistically significant compared to a baseline aging rate of 1 (p *<* 0.05, CI 2.54-3.02). To adjust for the bias introduced by the prediction error of the pre-RT scans in the SFCN model, as well as to show the heterogeneity of the slopes, figure 7/D shows the same regression slopes as figure 7/C with the intercepts removed. The smoothed average LOESS regression curve (red) shows an upward trend with the curve flattening as data points get sparser. All lines have a higher slope than normal aging, showing that the linear mixed effects model predicts every patient to age faster than normal, which indicates that all patients undergoing RT will show increased aging when measured with this framework. The narrow confidence bands show that the model has low uncertainty, especially up to 12 months.

### 3.4 Saliency maps

To determine which areas of the brain had the highest contribution in the SFCN model for determining brain age, a population average saliency map was created, shown in figure 8. A saliency map shows which voxels of an MRI scan contributed the most to the outcome. In figure 9 the thresholded saliency map is shown with a cortical atlas on top. The green crosshair emphasize a cluster of 753 voxels within the Heschl’s gyrus, which borders are shown in dark blue. The Heschl’s gyrus is associated with acoustic processing [34], indicating that the model weights could be translated to specific brain functions. Other examples of contributing anatomically well-defined areas are the brain stem and the middle cerebellar peduncle clusters with more than 1500 voxels.

**Fig. 8.**
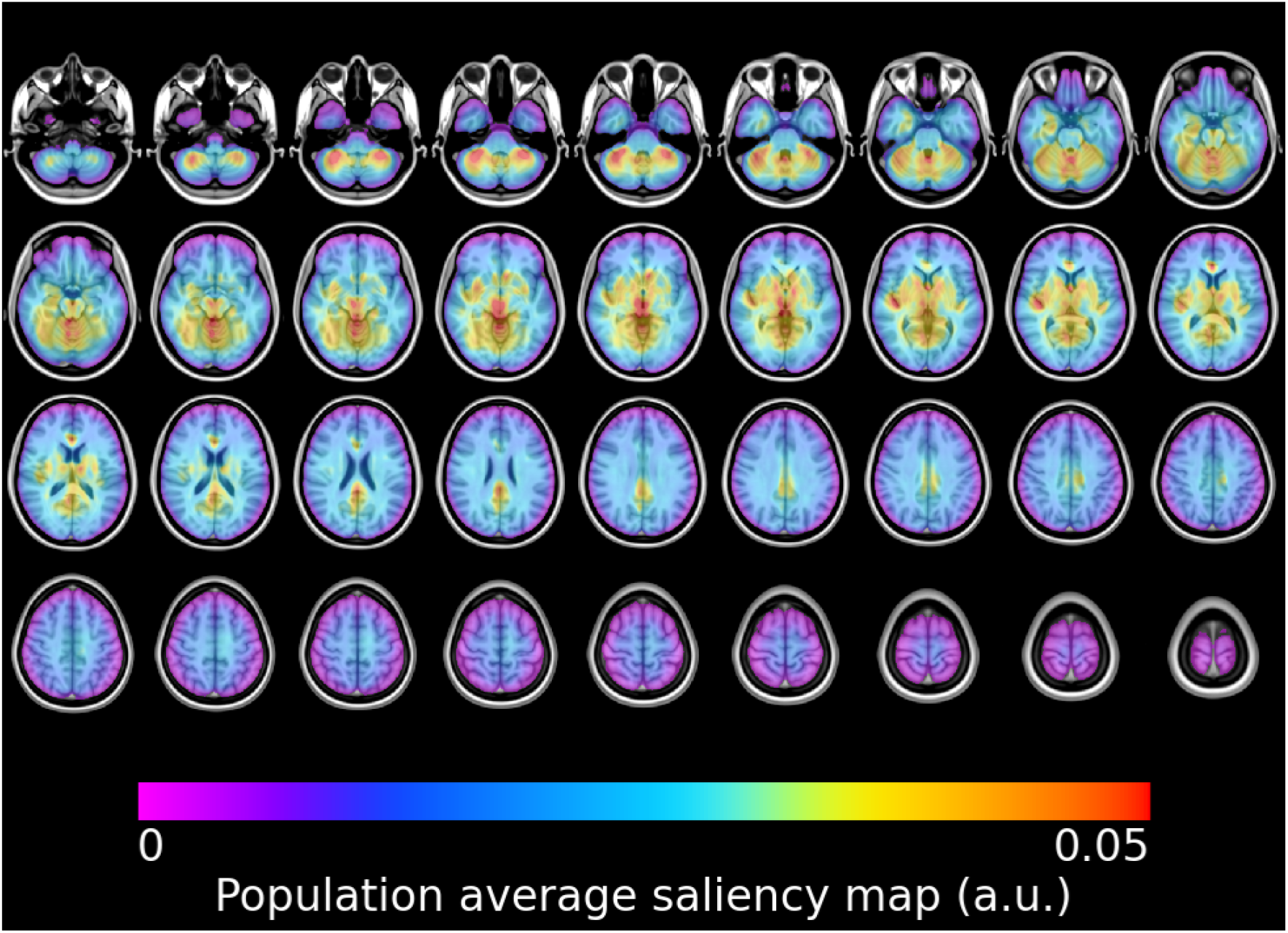
The population average saliency map, showing 27 layers of the brain, providing an overview of the weights of the model over the entire brain. Purple indicates the lowest weights, while red indicates the highest weights for the model.

**Fig. 9.**
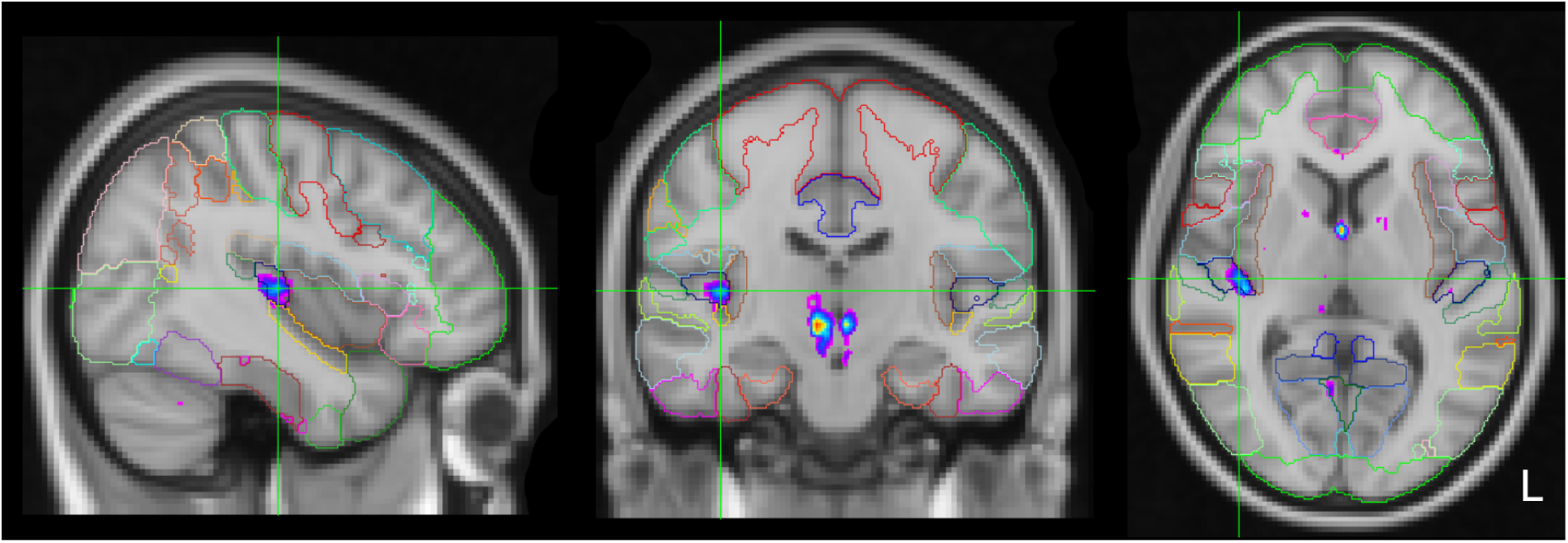
The population average saliency map with the background image of the MNI152 2mm template. The Harvard-Oxford cortical atlas [27] is overlaid, showing which brain regions contains the high-weighted clusters. One of which is the Heschl’s gyrus, under the green crosshair. The more warm (red) an area is coloured, the higher it is weighed by the model, with the purple being the least weighed.

Figure 10 shows the saliency maps from both the RT population and the healthy IXI population [16], to compare the difference in brain areas of importance between the two populations. The general structure of the saliency maps stay the same, although there are differences between the two, as seen in 10/C. The areas with a difference of more than 0.015 for the radiotherapy patients are the Heschl’s Gyrus and white matter in the cerebellum, while in the healthy cohort the brain stem had more contribution.

**Fig. 10.**
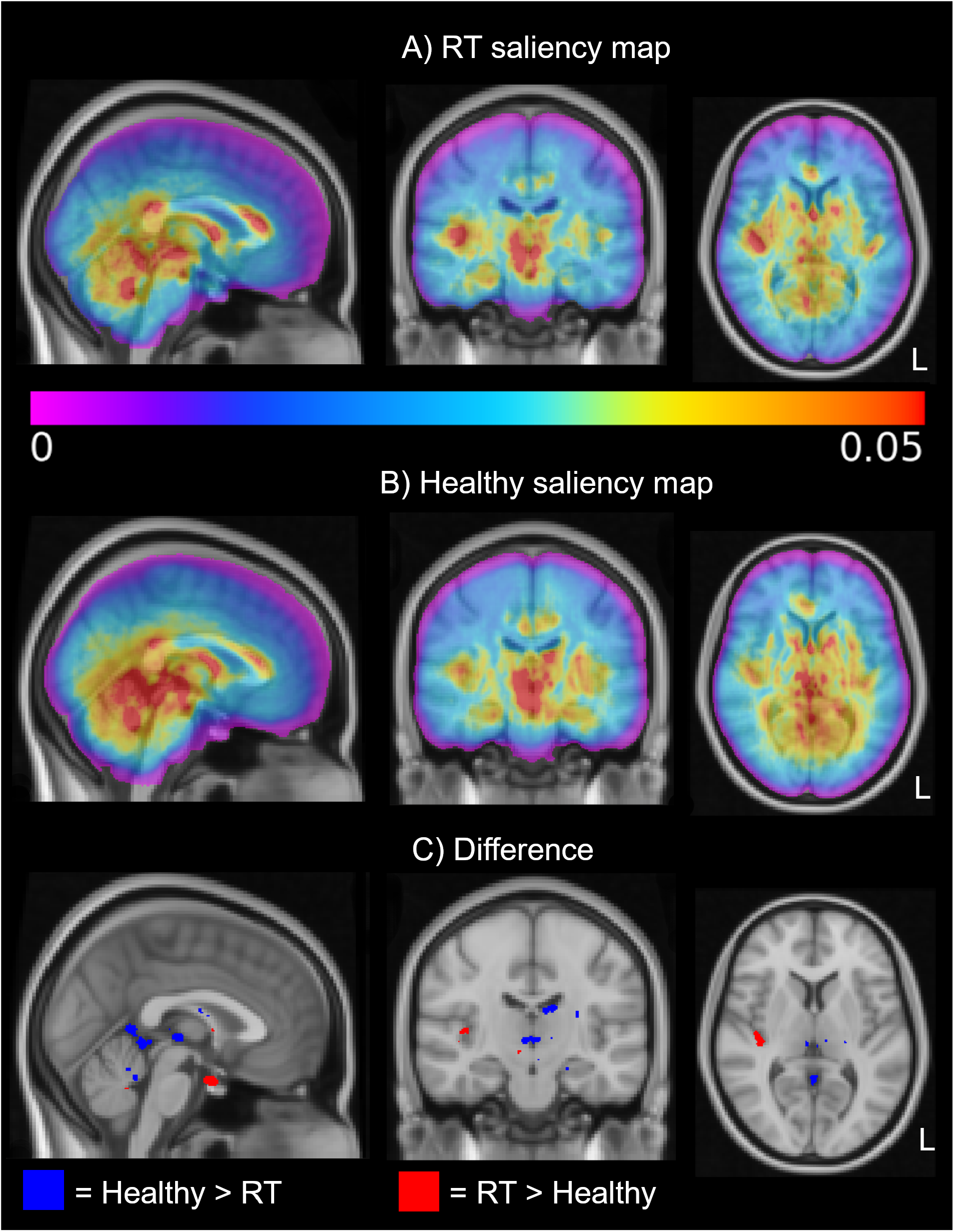
The population average saliency map compared to the population average saliency map from the IXI dataset. A) shows the RT patient group, while B) shows the healthy patient group. For the differences in C), red is a difference of 0.015 or more for the RT saliency map, blue is a 0.015 difference or more for the IXI saliency map.

In figure 11/A, the MRI scan of an example patient with a large resection cavity in the frontal part brain is shown, with the saliency map and the radiation dose on top. Since purple colour in the current colour scale indicates the lowest model weight, and no colour means zero weight, the model does not take the resection cavity into account for patient A. For patient B, the resection cavity does have a model weight, but the irradiated area shows a lower model weight than the rest of the brain.

**Fig. 11.**
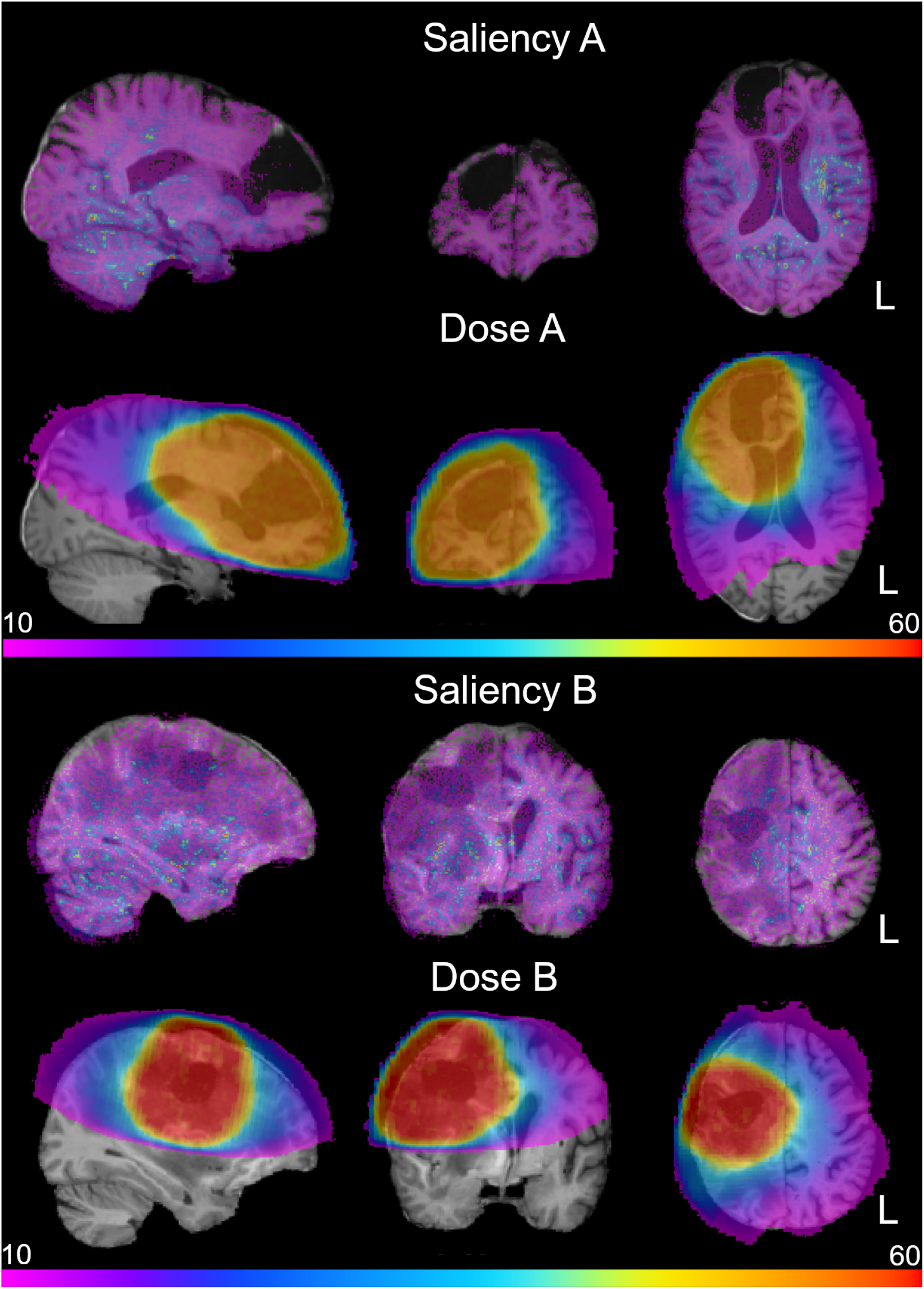
The saliency maps and dose for the brains of two example patients, both with a resection cavity in the brain. Red indicates a higher dose/model weight, while purple indicates a lower dose/model weight. The colour scale measures the amount of radiation in gray.

## 4 Discussion

In this study, we show that cranial RT has a remarkable effect on the brain, which we coined as radiation-induced accelerated brain aging. A deep learning model was able to quantify changes in the brain after RT. After RT, the brain showed a statistically significantly accelerated aging of 2.78 times the normal rate in 32 glioma patients (p *<* 0.05). Overall, these results imply that radiation changes the tissue of the brain, which manifests as accelerated aging. Based on studies investigating the effect of normal, healthy aging, an increased brain age after RT may also result in cognitive decline [15]. Normal appearing brain tissue should be spared as much as possible to avoid this radiation-induced accelerated aging, when irradiating the tumor-affected area.

The changes are quantified with an interpretable score using the entire brain without the potential bias or limitation of commonly used, pre-engineered features found in neuroimaging morphometry studies, such as cortical thickness [6] and volumetric measures [7], e.g.: hippocampus volume [35]. The population average saliency maps provide insights into the workings of the SFCN model. The areas weighed highest by the model are located in well defined anatomical areas of the brain, and may encompass certain functions of the brain, showing that the saliency maps could be of interest for further research into brain aging. The absence of model weights for brain abnormalities indicates that the model values the existing brain structure most, and bases the predictions on the existing tissue patterns. The regions that received the largest amount of radiation to fulfill the therapeutic needs have lower model weights in the saliency maps, indicating that brain tissue that received large amounts of radiation does not necessarily contribute more to the brain age prediction. While these results should be interpreted with caution due to the difference in saliency maps between models [36], they could provide a better understanding of brain aging.

Lastly, there are limitations to this study, starting with the baseline prediction error. The SFCN model has a relatively large prediction error for the pre-RT scans, which can be caused by several factors. All patients present some sort of abnormality, because of the tumor itself and treatment complication such as tissue scarring and oedema. Since the SFCN model was trained on healthy volunteers without such abnormalities, they are not represented in the prediction, which can affect the performance of the model. Another factor is the differences between scanners, as the deep learning model was trained on UK Biobank data, which uses different scanners and scanner parameters than our cohort. Despite the baseline error introduced by the pre-trained model, the trends shown by the model are clear, indicating that while the accuracy of the pre-RT predictions may not be perfect, the aggressive effect of RT acts through such model imperfections, measured by the increased aging rate. Furthermore, we assume that the baseline error established from the pre-RT scans applies systematically to all follow-up scans, therefore the measured upward trend in brain ages correspond to only-and-only to the RT-related tissue changes. We find this assumption permissive, as it is highly unlikely that the prediction errors largely collide with the measured effects, since such an error would have to have the same direction as the effect, while a random error is expected on a population level. For future work, BrainAGE may provide a novel way to quantify the effectiveness and damage caused by treatment by comparing patient outcomes. By selecting the treatment with reduced accelerated aging, damage to healthy brain tissue could be minimized.

## 5 Conclusion

In conclusion, in this work we show that patients who have undergone cranial RT experience brain tissue atrophy, which can be identified as radiation-induced accelerated aging using the BrainAGE method. Due to the lack of pre-engineered features, BrainAGE can be used to predict aging rates in a non-biased manner. Since the saliency maps indicate that there is an aging effect occurring in the healthy tissue, a global aging effect might be present for the entire brain. This indicates that the mere presence of RT will cause radiation-induced accelerated aging, potentially affecting patients’ QoL. By comparing the radiation-induced accelerated aging between patients and selected the treatment with the least amount of aging, damage to healthy tissue caused by the treatment may be reduced.

## Data Availability

Raw imaging patient data are not available.
The healthy data from the IXI data set are available online at https://brain-development.org/ixi-dataset/.
The healthy data from the MyConnectome data set are available online at https://openneuro.org/datasets/ds000031/versions/1.0.0.
The SFCN model is available online at https://github.com/ha-ha-ha-han/UKBiobank_deep_pretrain.

https://openneuro.org/datasets/ds000031/versions/1.0.0

https://github.com/ha-ha-ha-han/UKBiobank_deep_pretrain

## 6 Data sharing

MRI scans of the patients cannot be shared. The IXI data set can be found at [16]. The MyConnectome data set can be found at [37]. The SFCN model code can be found at [38].

## 7 Acknowledgements

The exact ages from patients have been replaced with age ranges to comply with medRxiv requirements.

## References

1. Chen, B., Dai, W., He, B., Zhang, H., Wang, X., Wang, Y., Zhang, Q.: Current Multistage Drug Delivery Systems Based on the Tumor Microenvironment. Theranostics. 7, 538–558 (2017).

2. Parhi, P., Mohanty, C., Sahoo, S.: Nanotechnology-based combinational drug delivery: an emerging approach for cancer therapy. Drug Discovery Today. 17, 1044–1052 (2012).

3. Mitchell, D., Fecci, P., Sampson, J.: Immunotherapy of malignant brain tumors. Immunological Reviews. 222, 70–100 (2008).

4. Fjell, A., Walhovd, K., Fennema-Notestine, C., McEvoy, L., Hagler, D., Holland, D., Brewer, J., Dale, A.: One-Year Brain Atrophy Evident in Healthy Aging. Journal of Neuroscience. 29, 15223–15231 (2009).

5. David, S., Mesri, H., Bodiut, V., Nagtegaal, S., Elhalawani, H., de Luca, A., Philippens, M., Viergever, M., Mohamed, A., Ding, Y., Chung, C., Fuller, C., Verhoeff, J., Leemans, A.: Dose-dependent degeneration of noncancerous brain tissue in post-radiotherapy patients: A diffusion tensor imaging study. (2019).

6. Nagtegaal, S., David, S., Snijders, T., Philippens, M., Leemans, A., Verhoeff, J.: Effect of radiation therapy on cerebral cortical thickness in glioma patients: Treatment-induced thinning of the healthy cortex. Neuro-Oncology Advances. 2, (2020).

7. Nagtegaal, S., David, S., van der Boog, A., Leemans, A., Verhoeff, J.: Changes in cortical thickness and volume after cranial radiation treatment: A systematic review. Radiotherapy and Oncology. 135, 33–42 (2019).

8. Franke, K., Ziegler, G., Klöppel, S., Gaser, C.: Estimating the age of healthy subjects from T1-weighted MRI scans using kernel methods: Exploring the influence of various parameters. NeuroImage. 50, 883–892 (2010).

9. Franke, K., Gaser, C.: Longitudinal Changes in Individual BrainAGE in Healthy Aging, Mild Cognitive Impairment, and Alzheimer’s Disease. GeroPsych. 25, 235–245 (2012).

10. Besteher, B., Gaser, C., Nenadić, I.: Machine-learning based brain age estimation in major depression showing no evidence of accelerated aging. Psychiatry Research: Neuroimaging. 290, 1–4 (2019).

11. Nenadić, I., Dietzek, M., Langbein, K., Sauer, H., Gaser, C.: BrainAGE score indicates accelerated brain aging in schizophrenia, but not bipolar disorder. Psychiatry Research: Neuroimaging. 266, 86–89 (2017).

12. Franke, K., Gaser, C.: Ten Years of BrainAGE as a Neuroimaging Biomarker of Brain Aging: What Insights Have We Gained?. Frontiers in Neurology. 10, (2019).

13. Makale, M.T., McDonald, C.R., Hattangadi-Gluth, J.A., Kesari, S.: Mechanisms of radiotherapy-associated cognitive disability in patients with brain tumours. Nature Reviews Neurology. 13, 52–64 (2016).

14. Tang, Y., Luo, D., Rong, X., Shi, X., Peng, Y.: Psychological disorders, cognitive dysfunction and quality of life in nasopharyngeal carcinoma patients with radiation-induced brain injury. PLoS ONE. 7, (2012).

15. MacDonald, M., Pike, G.: MRI of healthy brain aging: A review. NMR in Biomedicine. 34, (2021).

16. IXI Dataset – Brain Development, https://brain-development.org/ixi-dataset/.

17. Poldrack, R., Laumann, T., Koyejo, O., Gregory, B., Hover, A., Chen, M., Gorgolewski, K., Luci, J., Joo, S., Boyd, R., Hunicke-Smith, S., Simpson, Z., Caven, T., Sochat, V., Shine, J., Gordon, E., Snyder, A., Adeyemo, B., Petersen, S., Glahn, D., Reese Mckay, D., Curran, J., Göring, H., Carless, M., Blangero, J., Dougherty, R., Leemans, A., Handwerker, D., Frick, L., Marcotte, E., Mumford, J.: Long-term neural and physiological phenotyping of a single human. Nature Communications. 6, (2015).

18. Lutkenhoff, E., Rosenberg, M., Chiang, J., Zhang, K., Pickard, J., Owen, A., Monti, M.: Optimized Brain Extraction for Pathological Brains (optiBET). PLoS ONE. 9, e115551 (2014).

19. Jenkinson, M., Bannister, P., Brady, M., Smith, S.: Improved Optimization for the Robust and Accurate Linear Registration and Motion Correction of Brain Images. NeuroImage. 17, 825–841 (2002).

20. Jenkinson, M., Smith, S.: A global optimisation method for robust affine registration of brain images. Medical Image Analysis. 5, 143–156 (2001).

21. Peng, H., Gong, W., Beckmann, C., Vedaldi, A., Smith, S.: Accurate brain age prediction with lightweight deep neural networks. Medical Image Analysis. 68, 101871 (2021).

22. Woolrich, M.W., Jbabdi, S., Patenaude, B., Chappell, M., Makni, S., Behrens, T., Beckmann, C., Jenkinson, M., Smith, S.M.: Bayesian analysis of neuroimaging data in FSL. NeuroImage. 45, (2009).

23. Smith, S.: Fast robust automated brain extraction. Human Brain Mapping. 17, 143–155 (2002)

24. Allen, N., Sudlow, C., Peakman, T., Collins, R.: UK Biobank Data: Come and Get It. Science Translational Medicine. 6, 224ed4–224ed4 (2014).

25. Predicting Chronological Age from Structural Neuroimaging: The Predictive Analytics Competition 2019, https://www.frontiersin.org/research-topics/13501/predicting-chronological-age-from-structural-neuroimaging-the-predictive-analytics-competition-2019.

26. Python Software Foundation, Python Language Reference, version 3.85, http://www.python.org.

27. Desikan, R., Ségonne, F., Fischl, B., Quinn, B., Dickerson, B., Blacker, D., Buckner, R., Dale, A., Maguire, R., Hyman, B., Albert, M., Killiany, R.: An automated labeling system for subdividing the human cerebral cortex on MRI scans into gyral based regions of interest. NeuroImage. 31, 968–980 (2006).

28. Diedrichsen, J., Balsters, J., Flavell, J., Cussans, E., Ramnani, N.: A probabilistic MR atlas of the human cerebellum. NeuroImage. 47, S122 (2009).

29. Wakana, S., Caprihan, A., Panzenboeck, M., Fallon, J., Perry, M., Gollub, R., Hua, K., Zhang, J., Jiang, H., Dubey, P., Blitz, A., van Zijl, P., Mori, S.: Reproducibility of quantitative tractography methods applied to cerebral white matter. NeuroImage. 36, 630–644 (2007).

30. Warrington, S., Bryant, K., Khrapitchev, A., Sallet, J., Charquero-Ballester, M., Douaud, G., Jbabdi, S., Mars, R., Sotiropoulos, S.: XTRACT - Standardised protocols for automated tractography in the human and macaque brain. NeuroImage. 217, 116923 (2020).

31. RStudio Team: RStudio: Integrated Development for R. RStudio, https://www.scirp.org/reference/referencespapers.aspx?referenceid=2857579. (2020).

32. Bates, D. et al.: Fitting Linear Mixed-Effects Models Usinglme4. Journal of Statistical Software. 67, 1, (2015).

33. Jacoby, W.: Loess:: a nonparametric, graphical tool for depicting relationships between variables. Electoral Studies. 19, 577–613 (2000).

34. Warrier, C., Wong, P., Penhune, V., Zatorre, R., Parrish, T., Abrams, D., Kraus, N.: Relating Structure to Function: Heschl’s Gyrus and Acoustic Processing. Journal of Neuroscience. 29, 61–69 (2009).

35. Nagtegaal, S., David, S., Philippens, M., Snijders, T., Leemans, A., Verhoeff, J.: Dose-dependent volume loss in subcortical deep grey matter structures after cranial radiotherapy. Clinical and Translational Radiation Oncology. 26, 35–41 (2021).

36. Arun, N., Gaw, N., Singh, P., Chang, K., Aggarwal, M., Chen, B., Hoebel, K., Gupta, S., Patel, J., Gidwani, M., Adebayo, J., Li, M., Kalpathy-Cramer, J.: Assessing the Trustworthiness of Saliency Maps for Localizing Abnormalities in Medical Imaging. Radiology: Artificial Intelligence. 3, (2021).

37. Poldrack, R.: Myconnectome, https://openneuro.org/datasets/ds000031/versions/1.0.0.

38. Peng, H., Gong, W., Beckmann, C., Vedaldi, A., Smith, S.: GitHub - ha-ha-ha-han/UKBiobank deep pretrain: Pretrained neural networks for UK Biobank brain MRI images. SFCN, 3D-ResNet etc., https://github.com/ha-ha-ha-han/UKBiobankdeeppretrain.

